# Development and testing of a game-based digital intervention for working memory training in autism spectrum disorder

**DOI:** 10.1101/2020.04.23.20030494

**Authors:** Surbhit Wagle, Arka Ghosh, P Karthic, Akriti Ghosh, Tarana Pervaiz, Rashmi Kapoor, Koumudi Patil, Nitin Gupta

**Author notes:** Corresponding author, Postal address: Dr. Nitin Gupta, Department of Biological Sciences and Bioengineering, Indian Institute of Technology Kanpur, Kanpur 208016, Uttar Pradesh, India.

## Abstract

Autism spectrum disorder (ASD) is prevalent globally, yet it lacks cost-effective treatment approaches. Deficits in executive functions occur frequently in autism spectrum disorder and present a target for intervention. Here we report the design and development of five smartphone-based games for training working memory in autistic children. These open-source games, available free of cost to the community, were designed to match the behavioral preferences and sensorimotor abilities of autistic kids. We then conducted a preliminary trial to test the effectiveness of a month-long intervention using these games. Although we did not see a significant change in the working memory of all children with a month-long training, children who performed better on the games also showed more improvement in their working memory, suggesting that a longer intervention with the games might be useful in improving working memory. Using a Hindi translation of the Autism Treatment Evaluation Checklist (ATEC), we also tested the far transfer of the training in reducing autistic symptoms. We found no significant change in the autistic symptoms after the intervention. Further, there was no correlation between the change in the working memory and the change in the autistic symptoms.

## Introduction

Autism Spectrum disorder (ASD) is a neurodevelopmental disorder characterized by problems in communication, social interactions, idiosyncrasy, and repetitiveness in behaviors ^1^. According to WHO estimations, 1 in every 160 children worldwide is on the ASD ^2^. The country-specific prevalence rates are 1.70 % in the USA ^3^, 2.64% in South Korea ^4^, and 0.23% in India ^5^; lower rates in developing countries like India may partly be due to lack of awareness and diagnoses. Current treatments for ASD include early interventions like Applied Behavioral Analysis ^6,7^. These treatments are cost-and time-intensive^8,9^, and hence often limited in their access, particularly in developing countries. There is a pressing need for developing and testing new cost-effective treatment approaches for ASD.

Executive Functions (EFs) ─ cognitive processes such as working memory (WM), attention, cognitive flexibility, planning, set shifting ─ are essential for the day-to-day functioning of people. Individuals with neurodevelopmental disorders are likely to have some deficits in EFs, although the deficits may differ across the affected individuals ^10^. The commonalities between the symptoms of ASD and the symptoms of frontal lobe lesions led to the cognitive theory of executive dysfunction for explaining symptoms of ASD ^11^. WM impairments are related with learning disabilities, other EF deficits ^12^, and restricted and repetitive behavior^13,14,15^. One study found that high functioning ASD individuals made more between-search errors and employed a less strategic search compared to their typically developing (TD) peers ^15^. Several studies have reported that in ASD individuals verbal WM impairments increase with an increase in WM load and are absent when the load is minimum^16,17^. Similarly, for the visual-spatial WM, studies have reported significantly poorer performance of the ASD individuals compared to the TD control group in spatial working memory tasks ^18^. However, other studies have reported comparable performance in the two groups^19,20,21,22,23^. These inconsistencies might be due to the differences in the severity of ASD in the participants ^24^ and the type of WM (verbal or spatial) examined ^12^. A recent meta-review concluded that ASD individuals have deficits in verbal and spatial working memory and these deficits are present in individuals in a broad age range ^25^.

Effects of EF training for clinical and TD populations have been investigated previously^26,27,28^. Several studies have looked into the benefits of using computerized WM training for ADHD, learning disability, and TD populations^29,30^. These studies found significant near-transfer effect to WM, the trained EF, although the far-transfer to other EFs was less clear ^31^. Improvements in cognitive abilities in young children with ASD can concurrently show improvement in their social skills ^32^. This raises the possibility that WM training may reduce ASD symptoms, since studies have shown a link between WM and the repetitive and restricted behaviors in ASD^13,14,15^. Baltruschat et al. were able to show improvements in WM task performance of autistic individuals with the help of positive reinforcement^33,34^. They used items that were highly preferred by these individuals as the positive reinforcement. Also, the high performance was maintained at follow-up, when the reinforcement was removed^33,34^. This suggests opportunity to train WM using incentives that are preferable to ASD population.

Despite the evidence of WM deficits in ASD population and of positive results of WM training in similar disorders, very few studies have explored WM training as a potential intervention for ASD. In a recent study, effects of computerized working memory training or cognitive flexibility training, over 6 weeks, were compared against a mock training in ASD children ^35^. At post-training, both the experimental groups showed near-transfer improvements in their respective trained EFs. The working memory training showed marginal improvement in attention at post-training ^35^. The study, however, suffered from a relatively high number of dropouts. Here we explore the possibility whether gamified and mobile-based WM training can serve as a cost-effective and time-efficient intervention for ASD. We developed a suite of mobile-based games to train visuo-spatial WM in autistic children. Our aim was to develop simple and interesting games that are able to engage children with ASD and are freely available to all children. Although the games were based on English language, only the simplest words were used and the use of language was kept to a minimum, so that the games can be used by non-native English-speaker globally. To incentivize gameplay, our games were designed to align with the behavioral preferences of ASD children, and were improved using iterative user testing. Finally, we conducted a preliminary study to test the effectiveness of a short-term (30 minutes per day for one month) intervention using our games in improving WM and reducing autism severity.

## Methods

### Game development process

We sought to develop a suite of five games for providing working memory training to ASD children. The games were designed from scratch taking into account the behavioral preferences and the sensorimotor abilities of autistic children to incentivize and simplify the gameplay. It has been observed that autistic children prefer green and brown and show slight aversion towards the yellow color ^36^. Accordingly, we reduced the use of yellow in the games and instead made more game elements using the preferred colors. We kept navigation in the games simple to keep the game play accessible to most children: for instance, the games mostly required tapping of objects instead of dragging. Considering the short visual attention spans of autistic children, the games were designed to have single windows with very few elements on the screen to minimize distractions. An earlier study showed improvement in task engagement when the directives were sung instead of spoken ^37^. Therefore, we used song-like intonations in complimentary messages such as “well done” and “you are awesome”, which are provided when the child successfully completes different steps in the games. Lining up toys is one of the common repetitive behaviors observed in autistic children ^38^. We exploited this preference to motivate the kids by designing gameplays where the game objects get arranged in a line as the player progresses through the game. We further optimized our game development process based on the inputs from parents and caretakers of autistic children, and based on a survey of existing games (and their reviews) on the Google Play and the iTunes store. We iteratively refined the initial versions of the games by user testing with the children and by taking feedback from the therapists at the Amrita School for Special Children, Kanpur, India.

The games were programmed to run on smartphones or tablet devices with the Android Operating System. We targeted Android, as it is the most widely used mobile operating system globally. The games were developed using Unity 2018.3.5f1 game engine. The games are made freely available on the Play Store at this link: https://play.google.com/store/apps/details?id=org.TreadWill.WorkingMemoryGames&hl=enY. The source code of the games is also made freely available at this link: https://github.com/neuralsystems/working-memory-training-games.

### Participants

All participants were recruited from Amrita School for Special Children, Kanpur, India. 19 parents responded to our request for participation in this study. After initial screening to exclude kids with serious behavioral problems who could not use tablet devices, 14 children (mean age = 10.24, SD = 1.92; 11 males, 3 females) were included in the trial as participants. Thirteen out of the fourteen children were diagnosed with autism spectrum disorder by a physician or a therapist at the center using DSM-V criteria ^1^, while one had a diagnosis of Down’s syndrome with some symptoms of ASD□. Eleven participants were regular students in the special-needs school run by the center. The remaining three participants visited the center for behavioral therapy and one-to-one interactions with the teachers of the school. The students visited the center for an average of 5 days/week. The non-student participants visited the center in varying frequencies: of the three non-student participants, one child visited 5 days/week, and two children visited 2 days/week. All of the participating children received the same intervention.

### Outcome measures

To determine the effectiveness of our games, we evaluated the change after the training in the working memory and the far-transfer to the autistic symptoms. These two effects were examined using the measures described below.

#### Working Memory

To record changes in visuospatial working memory, Corsi-block tapping task^39^ was administered. The task apparatus consisted of nine black-colored blocks randomly placed on a stage. In a single trial of the task, an examiner tapped a sequence on the blocks and the participant was instructed to repeat the tapped sequence as soon as the examiner finished tapping. The participant was instructed to refrain from tapping until the examiner finished tapping. The task started with an initial sequence length of two blocks and had a maximum sequence length limit of nine blocks. We tried two trials for each sequence length and used a different sequence in each trial. If the participant failed to produce at least one of the two sequences of same length correctly, the task was ended. If the participant correctly reproduced the sequences on the two trials, the sequence length was increased by one. Two dependent parameters in the task were noted. The first parameter is the length of the longest correctly produced sequence, also known as the Block span, and is commonly used for measuring performance on this task. The other parameter is the product of the Block span and the total number of correctly produced sequences, also known as the Total score. The latter is considered more reliable and sensitive to minor changes in the task performance^39^. We have used both the parameters as the outcome measures of the task.

#### Far-transfer to autistic symptoms

The Autism Treatment Evaluation Checklist (ATEC) ^40^ is a popular tool to measure the effectiveness of an intervention for patients on the autism spectrum. The ATEC form consists of 77 items that cover four major categories of difficulties observed on the autism spectrum. These categories: (i) Speech/Language/Communication (14 items); (ii) Sociability (20 items); (iii) Sensory/Cognitive Awareness (18 items); (iv) Health/Physical/Behavior (25 items). The first three categories are rated on a 0-2 scale and the fourth category is rated on a 0-3 scale. Parents provided rating for each item, which resulted in a total score between 0 and 179. This total score was used as the outcome measure of the ATEC. Since most of the parents were more comfortable with the Hindi language, we provided a Hindi translation of the ATEC for assessment (available at https://github.com/neuralsystems/ATEC_Hindi).

To assess the outcome of the working memory training, the above-mentioned measures were taken before and after the intervention for all the participants. Both assessments were taken within one week of the start and the completion of the intervention.

### Intervention

During the training session, the participant was presented with a headphone-plugged tablet with the game suite installed in it. All training sessions were monitored by an experimenter. In a session, the child had the liberty to play any of the games in the suite. The child was allowed to switch between the games as many times as he or she wanted, although the experimenter monitoring the session verbally encouraged the child to avoid switching too many times and play a game long enough to understand the gameplay and learn from it. Each training session lasted for at most 30 minutes.

### Procedure

The study was approved by the Institute Ethics Committee of the Indian Institute of Technology Kanpur. Parents of the children provided written consent for participation in the study. Subsequently, the participant was administered pre-treatment Corsi task, by either an experimenter or the participant’s favorite teacher at the center. For each participant, the same person administered the task pre-intervention and post-intervention so that any difference due to familiarity with the task administrators can be avoided. The teachers received a tutorial about the task procedure before they administered the task to any of the participants. For all the participants, their parents filled the pre-and the post-training ATEC form. For 13 out 14 children, the pre-and the post-treatment ATEC form was filled by the same parent. For one child, the mother filled the post-intervention form due to long-term inaccessibility of the father who had filled the pre-intervention form. Participants who were students in the school were provided the training during their school hours, depending on their availability from other school activities. The non-student participants were provided the training during their visit to the center for the therapy session. The methods are performed in accordance with approved guidelines and regulations. As the study was conducted as a preliminary trial, it was not registered prospectively; it was retrospectively registered at clinicaltrials.gov (NCT04308915).

### Data analysis

Given the small sample size available for our study, we did not assume the data to be normally distributed and used nonparametric tests to assess the statistical significance of the results. The analyses were performed in MATLAB. We calculated the change in Block Span as the difference in its values at the post-intervention and the pre-intervention time points. Similarly, the change in the total scores of Corsi task was calculated. Non-parametric Wilcoxon signed-rank tests were used to determine if the median change among the 14 participants is significantly different from zero. To determine the significance of the change in autistic symptoms, Wilcoxon signed-rank test was applied to the changes in the ATEC scores. Spearman rho values were used to assess the correlations between the changes in different outcome measures. For determining statistical significance, we set the P-value threshold to 0.05.

## Results

### Development of games for working memory training

The design process described in the **Methods** resulted in five games for working memory training in ASD children.

#### 1. Basket game

The objective in the game is to drop colored fruits into baskets of matching color. The player must tap on a moving bubble, containing a colored fruit, only when the bubble is above the basket of the same color (**Figure 1**). An audio-visual animation, which autistic kids find rewarding, is presented if the fruit falls in the correct basket. After a few correct plays, the colored part of all the baskets is hidden, thus requiring the player to remember the colors of the baskets at different positions. As the user is able to perform well, the difficulty in the levels is increased by increasing the number of baskets and the number of fruits per basket. We also added an introductory level to the game, in which the user learns the idea of collecting fruit into the basket. Here, we show single basket and one static fruit in a bubble. Tapping on the bubble bursts it and the fruit is collected in the basket. After several correct trials, we add movement to the fruit such that it starts moving in a zigzag path, starting from the bottom left corner of the screen. For a few trials, the fruit moves until it reaches above the basket and stops there. In the last few trials of the introductory level, the fruit does not stop above the baskets but continues to move. Thus we gradually teach the kid the idea of collecting moving fruits into the basket. In this introductory level, we have removed other all the objects from the screen to keep the attention of the child on learning the objective of the game.

**Figure 1:**
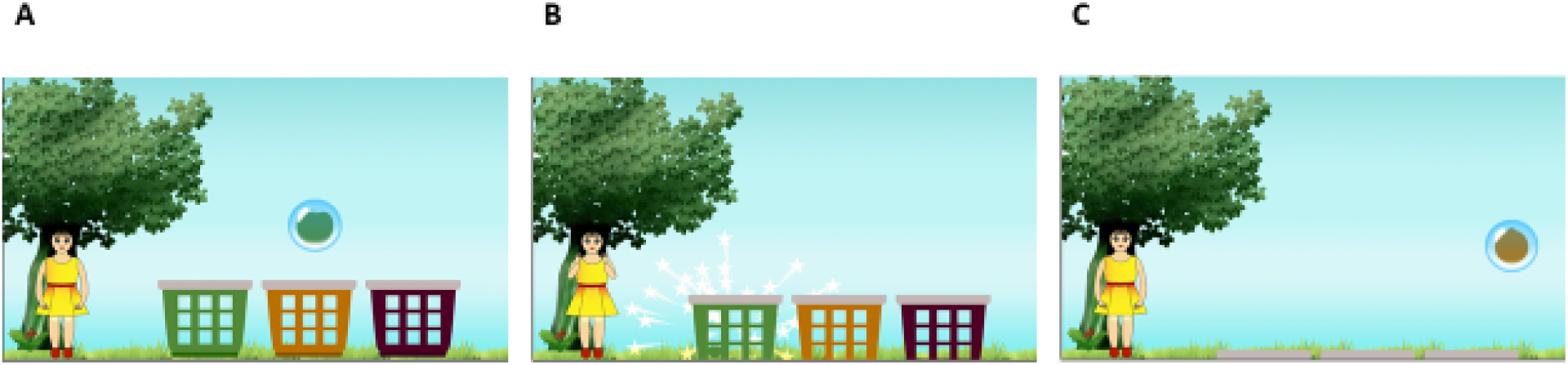
Basket Game gameplay. **(A)** A colored fruit is trapped inside a bubble, which moves in a zigzag path above the differently colored baskets. **(B)** The fruit falls in one of the baskets when the player bursts the bubble by tapping it. If the color of the fruit matches the color of the basket, a visual reinforcement is provided: the girl on the left claps and laughs, and a particle scattering effect is displayed. **(C)** As the game progresses, the baskets move down (and out of the screen) gradually such that eventually the colored parts of the baskets are completely hidden.

#### 2. Train Game

The objective in the game is to attach train wagons, each with specific shapes on its two ends, such that any two adjacent wagons have complementary shapes fitting snugly into each other (**Figure 2**). Correct play results in aligning of wagons and elongation of the train, which act as an incentive for the autistic children, who often have an inclination to arrange toys in lines. In the initial levels of the game, the shapes of the wagons are kept visible so that the player can understand the game play. As the player progresses to harder levels, the shapes of the wagons are hidden after being shown for a short duration, thus requiring the child to remember the shapes at different positions. Based on the feedback received in initial testing, we have also added some introductory levels in the game to teach the idea of matching complementary shapes (without wagons or other stimuli on screen) so that the children can slowly acquire the minimum skills required to play the game.

**Figure 2:**
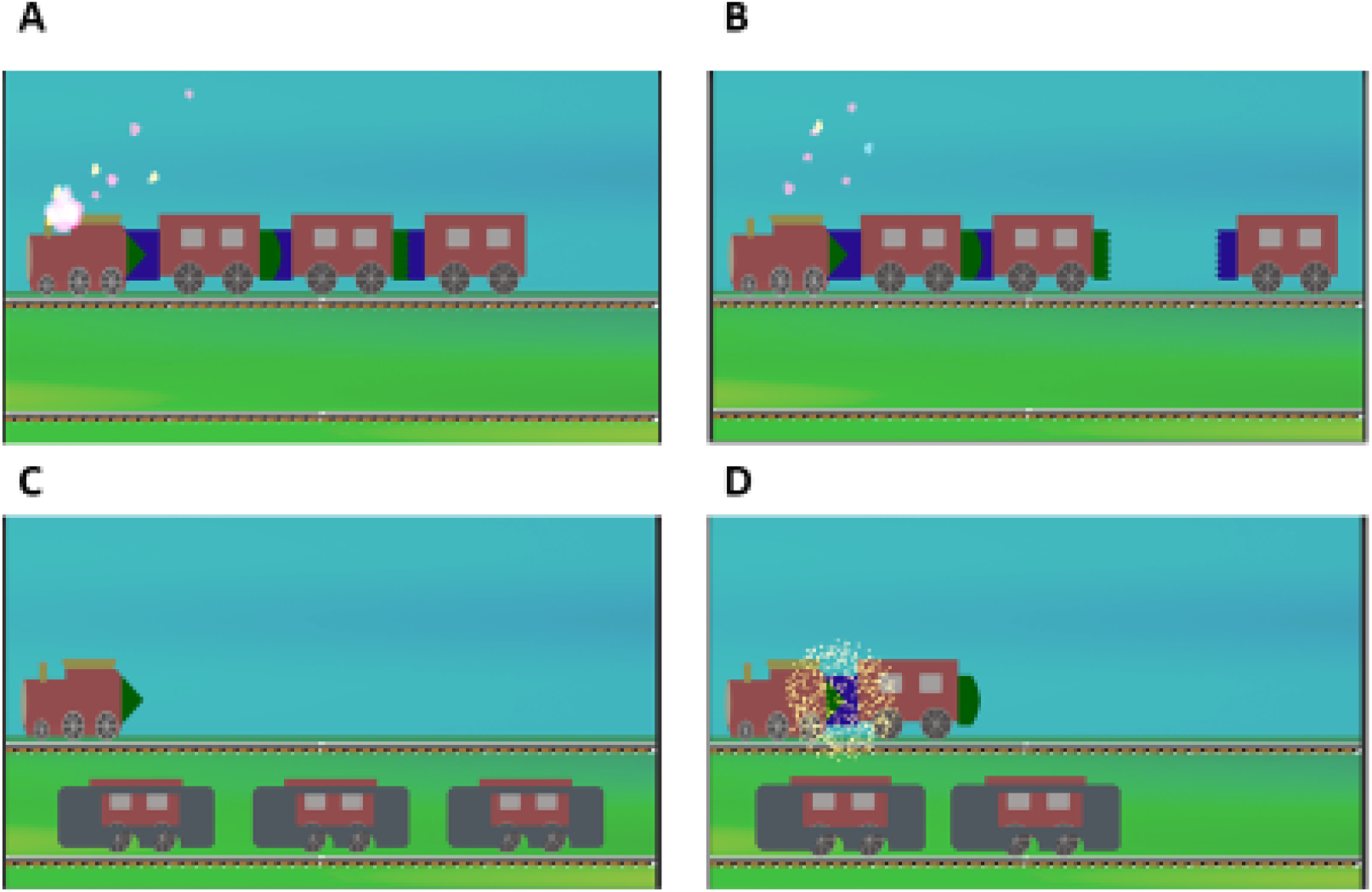
Train Game gameplay. **(A)** A train moves into the scene with smoke animation and sound. **(B)** Each wagon detaches from the train and moves to a random position on the rail track below. **(C)** After some delay, the shapes at the ends of the wagons are hidden by a cover. The player is then prompted to select the wagon that has the shape complementary to the last wagon on the train. **(D)** Selection of the correct wagon results in elongation of the train and is rewarded by an audio-visual feedback.

#### 3. Piano Game

The objective in the game is to hear and then replay a short sequence of musical notes on a virtual piano on the screen (**Figure 3**). This game capitalizes on the penchant of autistic children towards music. The note sequences are taken from popular songs such as “Baa Baa Black Sheep” and “Humpty Dumpty”. We have used visual animations to make it easy to see the pressed keys. A pleasant audio-visual animation is displayed when the player correctly repeats the sequence; the correctly played keys are also added to an elongating line on top to further motivate autistic players. We do not provide any feedback for incorrect play, because we observed during the testing phase that, for some children, any kind of audio or visual feedback (even if given after mistakes) was enjoyable and could encourage them to repeat mistakes during the game. When the child is able to perform well, we increase the difficulty of the game by increasing the length of the sequence and the number of trials per sequence length.

**Figure 3:**
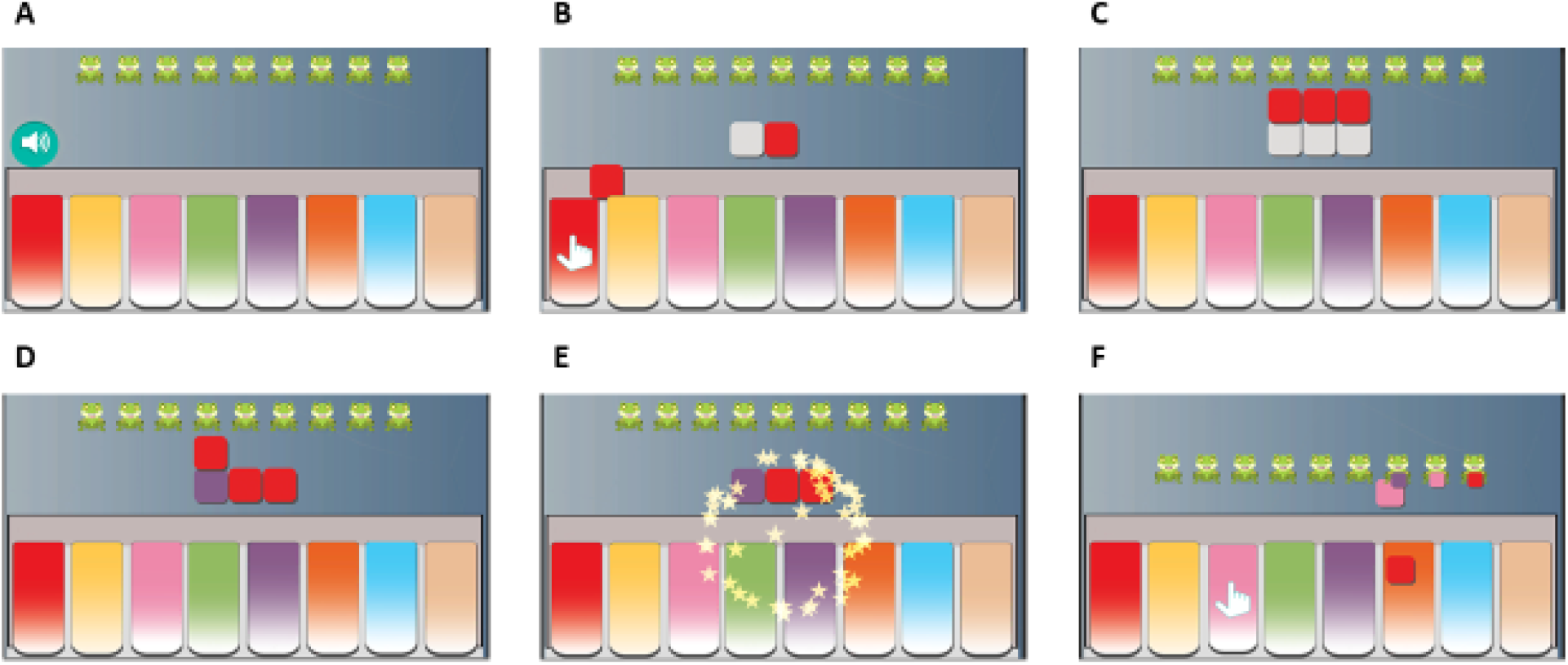
Piano Game gameplay. **(A)** The player clicks on the speaker icon to make the computer play a target sequence. **(B)** When the target sequence is played, tiles of the same colors as the played keys are generated and aligned above the keyboard. In addition, a visual cue (hand) indicates the pressed keys. After a delay, the colors of the tiles are hidden (color turns to white). Now, the player is prompted to repeat the target sequence using the keyboard. **(C)** As the player presses the keys, tiles corresponding to the pressed keys are arranged on top of the tiles of the target sequence. **(D)** When the player finishes repeating the sequence, the colors of the target tiles are uncovered, and the matches between the target and the player’s tiles are displayed. **(E)** An audio-visual reinforcement is provided if there is a complete match, and the play starts again with subsequent notes from the same song. The correctly played tiles also get added to a sequence of frogs at the top part of the screen. **(F)** At the end of a level, when the user has completed multiple sequences of a given length, the full song is played with animation. The user then progresses to the next level with longer sequences.

#### 4. Face Game

One of the common symptoms of ASD is avoiding eye contact and ignoring facial expressions during a social interaction (**Figure 4**). We developed this game to offer practice for identification of facial expressions along with working memory training. The objective in the game is to complete an empty face by adding facial components like hair, eyes, and mouth, such that the completed face matches a model face. For easier levels, we have used less detailed, cartoonish faces. As the player progresses to more difficult levels, more details are added to the face such that the faces look more real. After trying out an initial version of the games at the center, we noticed that many children were unable to understand that they have to match the components in order to complete the empty face. To overcome this difficulty, we added an introductory level in which the player learns to match the facial components individually without any face on the screen.

**Figure 4:**
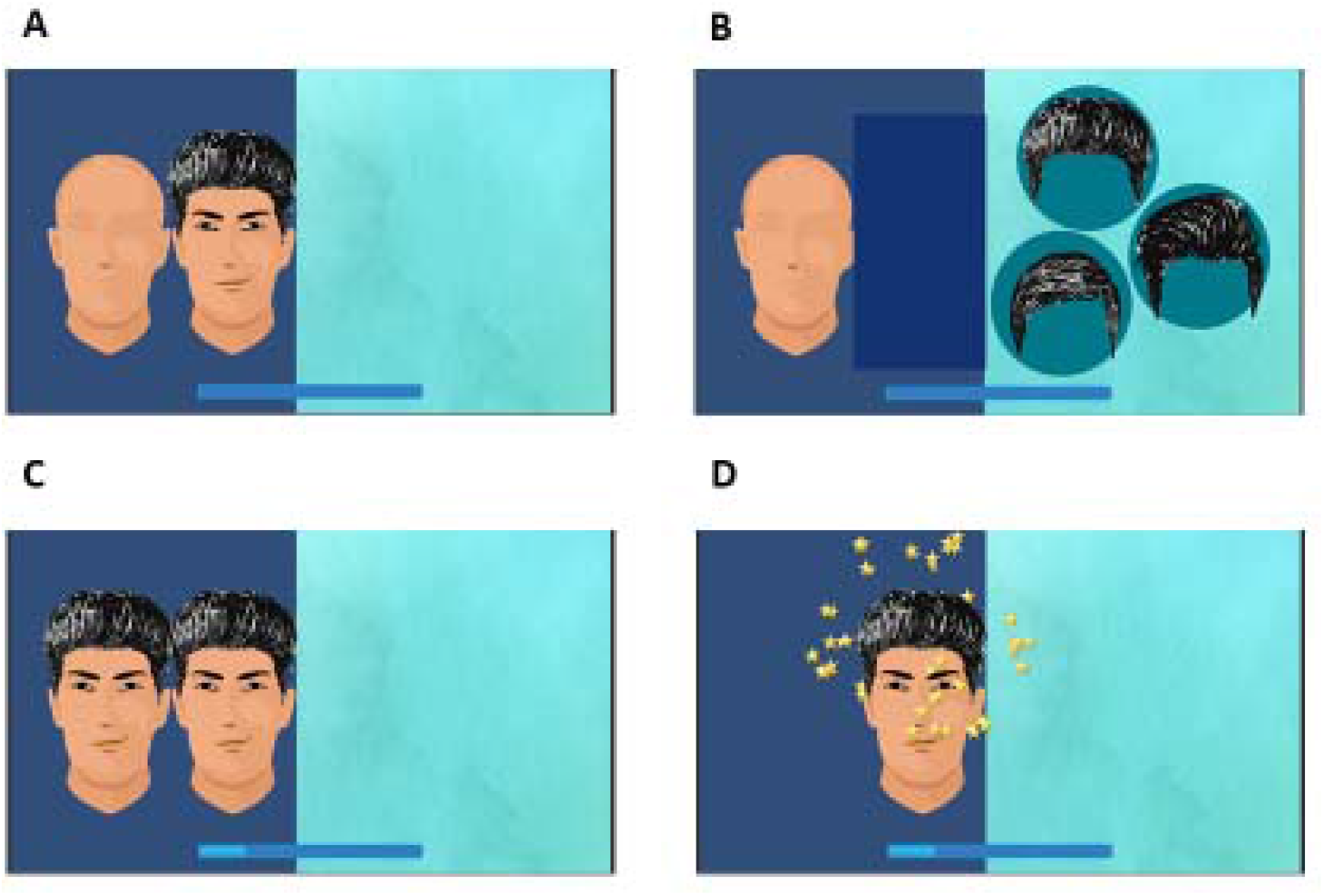
Face Game gameplay. **(A)** At the start of a level, a model face and an empty face lacking facial features are shown. **(B)** After a delay the model face is covered and for each facial feature the player is shown three options, from which the player selects one to add to the empty face, in attempt to make it the same as the model face. **(C)** Once all the components are added to the empty face, the model face is revealed and the similarity between the two is assessed. **(D)** A pleasant audio-visual feedback is given if the two faces match completely.

#### 5. Shape game

We designed this game especially for children who are at the lower end of the spectrum. The objective in the game is to match objects having the same shape, similar to the train game, but in this game the screen contains only simple shapes, some of which were hidden to challenge the working memory (**Figure 5**). To keep the gameplay simple, no other objects were added on the screen. For making the higher levels challenging, we increase the number of objects and also shuffle their positions after a correct match.

**Figure 5:**
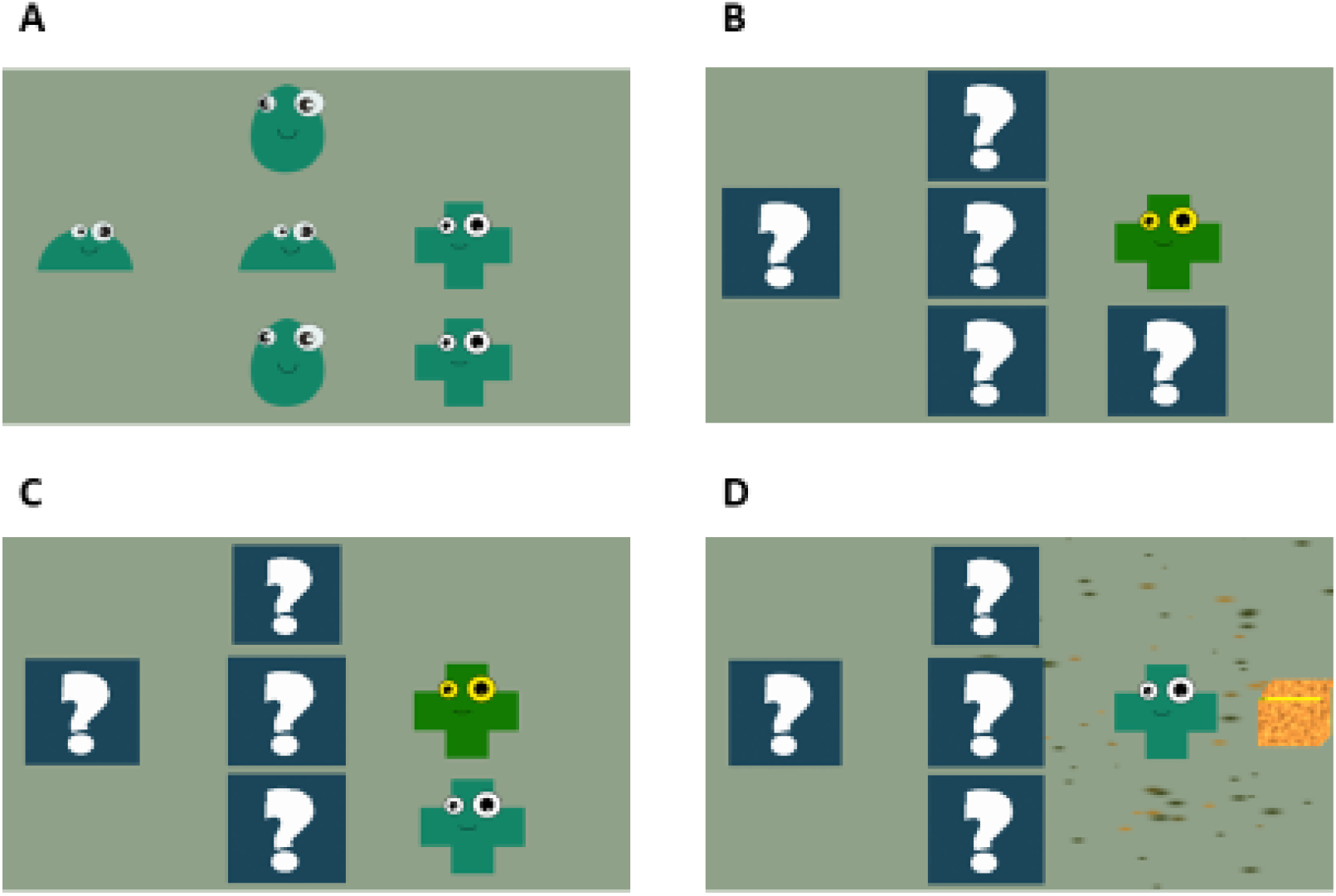
Shape Game gameplay. **(A)** Objects with different shapes (two of each shape) are shown at random locations on the screen. **(B)** The player taps on one object, after which all other objects are hidden. **(C)** The player needs to find the second object with the same shape by tapping one of the hidden objects. **(D)** If the player finds the correct object, a rewarding audio-visual feedback is provided. If the selected object is wrong, all the shapes are revealed, and the play starts over again.

In all the games, we have used a consistent wobbling animation to indicate which objects on the screen are supposed to be tapped when starting the game. For example, in the basket game, the bubble wobbles when it is moving in a zig-zag path to indicate that it should be tapped (the player has to use working memory to decide when to tap it). In each game, there are multiple difficulty levels, which differed in the number of objects (visible or hidden) in the game and therefore the working memory load. If the number of errors at the current level goes beyond a certain threshold – indicating that the player is finding the game difficult – the level in the game is automatically lowered by 1 step to reduce the difficulty. On the other hand, if the player performs well at the current level, the level is increased by 1 step to increase the difficulty. This adaptive level-adjustment allows maintaining the difficulty in each game at the child’s current competence level as the child progresses through the games, and prevents frustration or boredom that may be caused by very high or very low difficulty.

### Outcome measures

The means and SDs of the 14 participating children at the pre-intervention and the post-intervention time points are presented in **Table 1** and **Figures 6 & 7**.

**Table 1:** Mean and standard deviation values of the outcome measures before and after the training.

| Measures | Pre-intervention |  | Post-intervention |  |
| --- | --- | --- | --- | --- |
|  | Mean | SD | Mean | SD |
| Corsi task: Block span | 2.64 | 1.44 | 2.64 | 1.73 |
| Corsi task: Total score | 9.57 | 11.67 | 12.00 | 14.59 |
| ATEC: Total score | 78.57 | 19.62 | 72.14 | 24.22 |
| ATEC: Speech/Language/Communication | 15.35 | 6.35 | 14.14 | 7.22 |
| ATEC: Sociability | 18.35 | 4.70 | 16.07 | 5.22 |
| ATEC: Sensory/Cognitive Awareness | 17.71 | 7.17 | 17.21 | 7.74 |
| ATEC: Health/Physical/Behavior | 27.14 | 10.25 | 24.71 | 10.04 |

**Figure 6:**
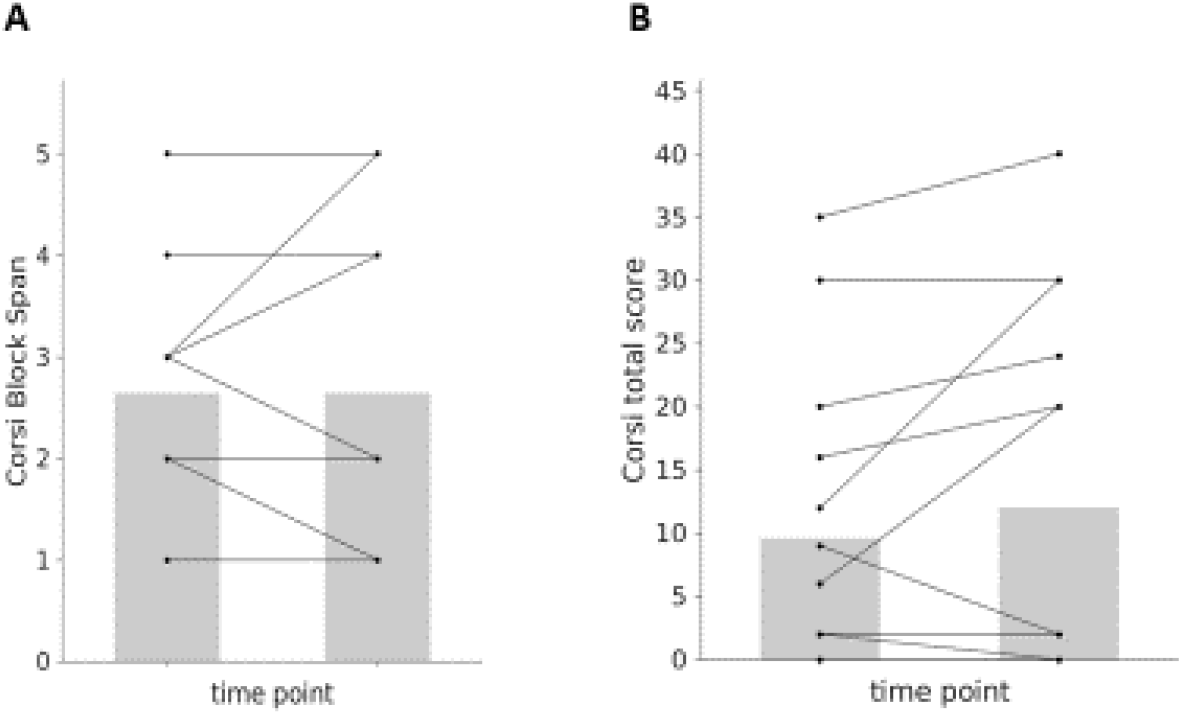
Short-term intervention with the games did not improve working memory. The graphs show pairwise comparisons between the pre-intervention and the post-intervention measures of working memory: The Corsi block span **(A)** and the Corsi total score **(B)**. No significant change was observed with either measure. Both plots have N=14 pairs of points, each corresponding to a subject, but some lines are overlapping.

**Figure 7:**
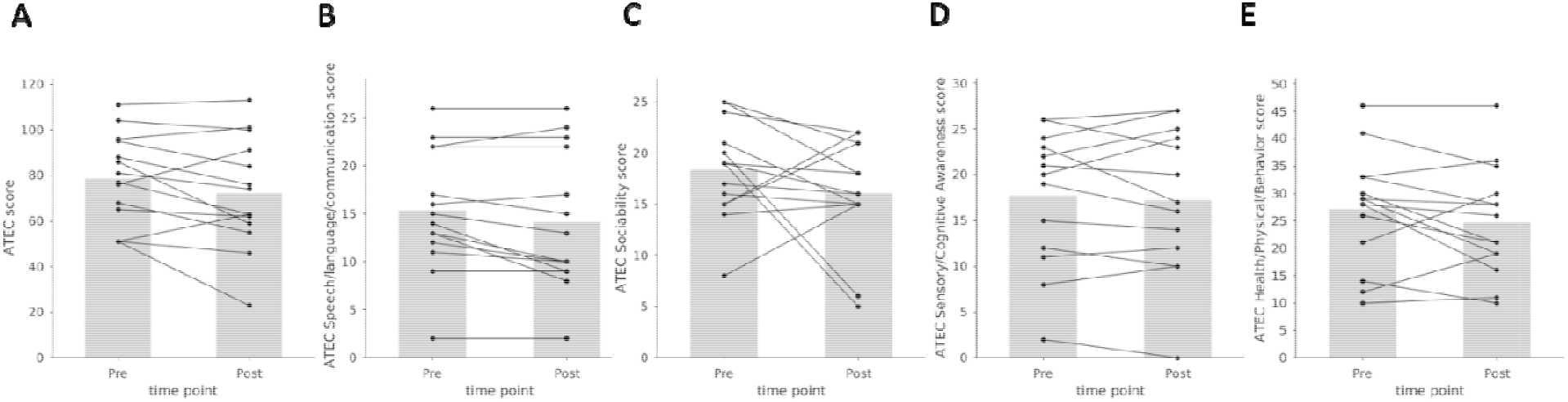
Short-term intervention with the games did not reduce autistic symptoms. The graphs show pairwise comparisons between the pre-intervention and the post-intervention measures of autistic symptoms measured using ATEC total score **(A)**, or the four sub-categories of ATEC, namely Speech/Language/Communication **(B)**, Sociability **(C)**, Sensory/Cognitive Awareness **(D)**, and Health/Physical/Behavior (**E**). No significant change was observed in any case.

No change (W = 7.5, n = 14, P = 1) was observed in the mean block spans between the pre and post intervention conditions (**Figure 6A**). Similarly, no significant change was observed in the Corsi total score (W = 27, n = 14, P = 0.22; **Figure 6B**). The ATEC score did not show a significant reduction after the intervention (W = 26, P = 0.10; **Figure 7A**). We further checked the change in each of the four subcategories of ATEC, and found that none of them showed a significant change: Speech/Language/Communication (W = 6, P = .06, **Figure 7B**); Sociability (W = 33, P = 0.24, **Figure 7C**); Sensory/Cognitive awareness (W = 44, P = 0.61, **Figure 7D**); Health/Physical/Behavior (W = 25, P = 0.16, **Figure 7E**). In summary, the children in our study did not gain significantly from the month-long game-based training in improving working memory or in reducing autistic symptoms.

Next we checked if there was any correlation between how well a child performed on the games and his or her improvements in the outcome measures. We noticed that the performances on different games were positively correlated (**Table 2**); in other words, kids who reached higher levels in one game also usually reached higher levels in other games. This was expected as all games were designed to test working memory using different gameplays. Therefore, instead of analyzing the performance on each game separately, we used the sum of the maximum levels in all five games achieved by a child as a measure of the performance on the games, henceforth referred to as the ‘total game performance’.

**Table 2.** Performances on the five games were correlated. Each value indicates the Spearman correlation and the corresponding P-value (in parenthesis) between maximum levels reached in a pair of games.

|  | Basket | Piano | Shape | Train |
| --- | --- | --- | --- | --- |
| Piano | 0.83 (0.00025) |  |  |  |
| Shape | 0.69 (0.0059) | 0.65 (0.012) |  |  |
| Train | 0.86 (0.000085) | 0.77 (0.0012) | 0.74 (0.0027) |  |
| Face | 0.38 (0.17) | 0.45 (0.10) | 0.76 (0.0017) | 0.55 (0.043) |

The total game performance showed a significant positive correlation with the change in the Corsi total score (Spearman rho = 0.68, P = 0.0071, n = 14; **Figure 8A**). A similar positive correlation was seen between the total game performance and the change in the Corsi Block Span (Spearman rho = 0.55, P = 0.04, n = 14). This finding suggests that although there was no significant increase in the working memory of all children as a group after the month-long intervention, the children who performed well on the games were more likely to show an increase in their working memory. This raises the possibility that a longer training on our games might be helpful in improving the working memory.

**Figure 8:**
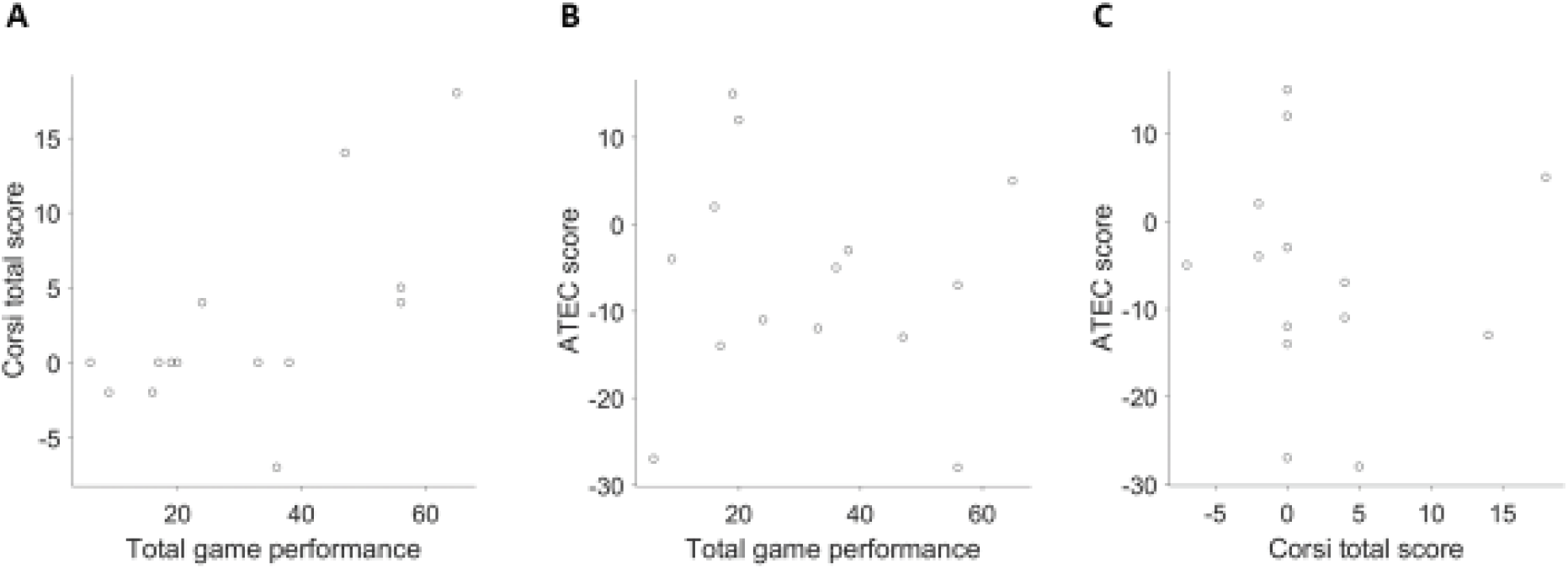
Performance on the games relates to improvements in working memory but not in autistic symptoms. **(A)** Scatter plot shows a positive correlation between the total game performance (the sum of maximum levels reached in different games) and the change in the Corsi total score with the intervention. **(B)** No correlation was seen between the total game performance and the change in the ATEC score. **(C)** Similarly, no correlation was observed between the change in the ATEC score and the change in the Corsi total score.

The total game performance was not correlated with the change in the ATEC score (Spearman rho = −0.03, P = 0.90; **Figure 8B**). Thus, the total game performance was correlated only with the working memory improvement but not with improvement in autistic symptoms. Consistent with this observation, we also found that there was no significant correlation between the change in the ATEC score and the change in the Corsi total score (Spearman rho = −0.25, P = 0.38; **Figure 8C**) or the Corsi Block Span (Spearman rho = −0.12, P = 0.68). Therefore, our limited month-long trial does not provide any evidence that short-term smartphone-based spatial working memory training is an effective intervention for reducing autistic symptoms.

## Discussion

In this study, we have developed mobile-based games for training working memory in autistic children. These games are well suited for children with neurodevelopmental disorders as they are designed keeping in mind the preferences of the target population and by doing iterative user testing during the development. The games are adaptive such that the difficulty level increases when the participant performs well and decreases when the participant finds the game difficult. In most of the games, we have added a few introductory levels to help the children understand the game idea in an intuitive and engaging manner. The introductory levels contain less number of elements than the main games to reduce distractions; the goals in the introductory levels are also simpler than the main games. The games are designed to require only simple interactions with the screen, such as tapping and keystrokes, which are easy enough for the autistic children ^41^; more complex interactions such as dragging are not used. This allows the children with problems in fine motor skills to play our games. The use of language was kept to a minimum, with only very simple English words used, so that the games can be used by children globally (in our study, children in the Hindi-speaking region of northern India were able to play the games).

We conducted a preliminary trial to evaluate the efficacy of short-term (one month) training with our games in improving the working memory and reducing autism severity. As the outcome of the trial, we measured changes in the performance of the participants on Corsi Block Tapping task and the rating on the ATEC provided by their parents. Overall, the participants did not show a significant improvement in their working memory or autistic symptoms after the short-term training. Our intervention was designed to be brief: both in the training intensity (up to 30 minutes per day) and the overall duration (one month), which was relatively low as compared to the 6-week or the 1-year durations used in the previous studies^27,42^. We found that the children who performed well on our games were more likely to improve their working memory, but these gains did not extend to autistic symptoms; further, there was no correlation between the improvement in the working memory and the improvement in the autistic symptoms. As the role of working memory in ASD remains debated, the results of various interventions such as ours contribute to the resolution of the debate, and from this standpoint, negative results are as informative as positive results. It is also important to look at the results of interventions from different cultural settings.

The sample size used in our trial was small (n = 14), as the trial was conducted as a pilot to estimate the effect sizes for our newly developed games, before a larger trial could be planned. The previous studies of similar kinds^26,27,42^ had larger sample sizes. The Corsi Block Spans (mean of 2.6) for the study participants were relatively low at pre-intervention condition than reported in previous studies^18,42,43^. This suggests that our participants had severe deficiency in the working memory, which combined with the short duration of the intervention might have been responsible for the lack of improvement after the training. It might be worthwhile to study in a larger trial whether the games can be useful for children with mild to moderate deficiency in working memory.

We have made the games freely available on the Android Play Store. We expect them to be of help to the community of researchers and therapists working with kids affected by ASD and other neurodevelopmental disorders. We have also released the source-code free of any restrictions so that other researchers can further build upon the framework of these games to create new training paradigms.

## Data Availability

The Game developed in this study is made available at the Google Play store. The source code of the games and the Hindi-version of the Autism Treatment Evaluation Checklist used in this study are available on GitHub.

https://play.google.com/store/apps/details?id=org.TreadWill.WorkingMemoryGames&hl=en

https://github.com/neuralsystems/working-memory-training-games

https://github.com/neuralsystems/ATEC_Hindi

## Acknowledgements

We thank Afifa Abdul Kader, Akanksha Sahu, Komal Meena, and Swastika Tandon for their help in designing, developing, and testing the games. We thank Braj Bhushan and Subramaniam Ganesh for helpful advice. We thank members of the Gupta laboratory for feedback on the games and the manuscript. NG is supported by the Wellcome Trust/DBT India Alliance Fellowship [grant number IA/I/15/2/502091].

## Author contributions

SW, Arka G, RK, KP and NG designed research; SW, PK, Akriti G and TP performed research; SW, Arka G and NG analyzed data; SW and NG wrote the paper with inputs from all co-authors.

## Competing Interests

The authors declare no competing interests.

## Notes

### Competing Interest Statement

The authors have declared no competing interest.

### Clinical Trial

We conducted a preliminary trial to evaluate the efficacy of a short-term (one month) training with our intervention. Our plan was to estimate the effect size for our newly developed intervention before conducting a large scale trial. Accordingly, the preliminary trial was not registered prospectively. It was retrospectively registered at clinicaltrials.gov (NCT04308915).

### Clinical Protocols

https://clinicaltrials.gov/ct2/show/NCT04308915

## References

1. American Psychiatric Association. Diagnostic and Statistical Manual of Mental Disorders. (American Psychiatric Associationx, 2013). doi:10.1176/appi.books.9780890425596.

2. Elsabbagh, M. et al.. Global Prevalence of Autism and Other Pervasive Developmental Disorders. Autism Res. 5, 160–179 (2012).

3. Baio, J. et al.. Prevalence of Autism Spectrum Disorder Among Children Aged 8 Years — Autism and Developmental Disabilities Monitoring Network, 11 Sites, United States, 2014. MMWR. Surveill. Summ. 67, 1–23 (2018).

4. Leventhal, B. L. et al.. Prevalence of Autism Spectrum Disorders in a Total Population Sample. Am. J. Psychiatry 168, 904–912 (2011).

5. Rudra, A. et al.. Prevalence of autism spectrum disorder and autistic symptoms in a school-based cohort of children in Kolkata, India. Autism Res. 10, 1597–1605 (2017).

6. Lovaas, O. I. Behavioral treatment and normal educational and intellectual functioning in young autistic children. J. Consult. Clin. Psychol. 55, 3–9 (1987).

7. Peters-Scheffer, N., Didden, R., Korzilius, H. & Sturmey, P. A meta-analytic study on the effectiveness of comprehensive ABA-based early intervention programs for children with Autism Spectrum Disorders. Res. Autism Spectr. Disord. 5, 60–69 (2011).

8. Peters-Scheffer, N., Didden, R., Korzilius, H. & Matson, J. Cost comparison of early intensive behavioral intervention and treatment as usual for children with autism spectrum disorder in the Netherlands. Res. Dev. Disabil. 33, 1763–1772 (2012).

9. Horlin, C., Falkmer, M., Parsons, R., Albrecht, M. A. & Falkmer, T. The cost of autism spectrum disorders. PLoS One 9, (2014).

10. Goldstein, S. & Naglieri, J. A. Handbook of executive functioning. Handbook of Executive Functioning (2014).

11. Damasio, A. R. & Maurer, R. G. A Neurological Model for Childhood Autism. Arch. Neurol. 35, 777–786 (1978).

12. Kercood, S., Grskovic, J. A., Banda, D. & Begeske, J. Working memory and autism: A review of literature. Res. Autism Spectr. Disord. 8, 1316–1332 (2014).

13. Landa, R. J. & Goldberg, M. C. Language, social, and executive functions in high functioning autism: A continuum of performance. J. Autism Dev. Disord. 35, 557–573 (2005).

14. Lopez, B. R., Lincoln, A. J., Ozonoff, S. & Lai, Z. Examining the relationship between executive functions and restricted, repetitive symptoms of Autistic Disorder. J. Autism Dev. Disord. 35, 445–460 (2005).

15. Sachse, M. et al.. Executive and Visuo-motor Function in Adolescents and Adults with Autism Spectrum Disorder. J. Autism Dev. Disord. 43, 1222–1235 (2013).

16. Williams, D. L., Goldstein, G., Carpenter, P. A. & Minshew, N. J. Verbal and Spatial Working Memory in Autism. J. Autism Dev. Disord. 35, 747–756 (2005).

17. Cui, J., Gao, D., Chen, Y., Zou, X. & Wang, Y. Working Memory in Early-School-Age Children with Asperger’s Syndrome. J. Autism Dev. Disord. 40, 958–967 (2010).

18. Verte, S., Geurts, H. M., Roeyers, H., Oosterlaan, J. & Sergeant, J. a. Executive functioning in children with autism and Tourette syndrome. Dev. Psychopathol. 17, 415–445 (2005).

19. Ozonoff, S. & Strayer, D. L. Further evidence of intact working memory in autism. J. Autism Dev. Disord. 31, 257–63 (2001).

20. Nakahachi, T. et al.. Discrepancy of performance among working memory-related tasks in autism spectrum disorders was caused by task characteristics, apart from working memory, which could interfere with task execution. Psychiatry Clin. Neurosci. 60, 312–318 (2006).

21. Gonzalez-Gadea, M. L. et al.. Cognitive variability in adults with ADHD and AS: Disentangling the roles of executive functions and social cognition. Res. Dev. Disabil. 34, 817–830 (2013).

22. Koshino, H. et al.. fMRI Investigation of Working Memory for Faces in Autism: Visual Coding and Underconnectivity with Frontal Areas. Cereb. Cortex 18, 289–300 (2008).

23. Geurts, H. M. et al.. How specific are executive functioning deficits in attention deficit hyperactivity disorder and autism? J. Child Psychol. Psychiatry Allied Discip. 45, 836–854 (2004).

24. Steele, S. D., Minshew, N. J., Luna, B. & Sweeney, J. A. Spatial working memory deficits in autism. J. Autism Dev. Disord. 37, 605–612 (2007).

25. Wang, Y. et al.. A Meta-Analysis of Working Memory Impairments in Autism Spectrum Disorders. Neuropsychol. Rev. 27, 46–61 (2017).

26. Fisher, N. & Happé, F. A Training Study of Theory of Mind and Executive Function in Children with Autistic Spectrum Disorders. J. Autism Dev. Disord. 35, 757–771 (2005).

27. Kenworthy, L. et al.. Randomized controlled effectiveness trial of executive function intervention for children on the autism spectrum. J. Child Psychol. Psychiatry. 55, 374–83 (2014).

28. Baltruschat, L. et al.. Addressing working memory in children with autism through behavioral intervention. Res. Autism Spectr. Disord. 5, 267–276 (2011).

29. Peijnenborgh, J. C. A. W., Hurks, P. M., Aldenkamp, A. P., Vles, J. S. H. & Hendriksen, J. G. M. Efficacy of working memory training in children and adolescents with learning disabilities: A review study and meta-analysis. Neuropsychol. Rehabil. 26, 645–672 (2016).

30. Shipstead, Z., Hicks, K. L. & Engle, R. W. Cogmed working memory training: Does the evidence support the claims? J. Appl. Res. Mem. Cogn. 1, 185–193 (2012).

31. Melby-Lervåg, M. & Hulme, C. Is working memory training effective? A meta-analytic review. Dev. Psychol. 49, 270–291 (2013).

32. McGovern, C. W. & Sigman, M. Continuity and change from early childhood to adolescence in autism. J. Child Psychol. Psychiatry 46, 401–408 (2005).

33. Baltruschat, L. et al.. Addressing working memory in children with autism through behavioral intervention. Res. Autism Spectr. Disord. 5, 267–276 (2011).

34. Baltruschat, L. et al.. Further analysis of the effects of positive reinforcement on working memory in children with autism. Res. Autism Spectr. Disord. 5, 855–863 (2011).

35. de Vries, M., Prins, P. J. M., Schmand, B. A. & Geurts, H. M. Working memory and cognitive flexibility-training for children with an autism spectrum disorder: a randomized controlled trial. J. Child Psychol. Psychiatry 56, 566–576 (2015).

36. Grandgeorge, M. & Masataka, N. Atypical Color Preference in Children with Autism Spectrum Disorder. Front. Psychol. 7, 1976 (2016).

37. Paul, A. et al.. The effect of sung speech on socio-communicative responsiveness in children with autism spectrum disorders. Front. Hum. Neurosci. 9, 555 (2015).

38. Kim, S. H. & Lord, C. Restricted and repetitive behaviors in toddlers and preschoolers with autism spectrum disorders based on the Autism Diagnostic Observation Schedule (ADOS). Autism Res. 3, 162–173 (2010).

39. Kessels, R. P. C., Van Zandvoort, M. J. E., Postma, A., Kappelle, L. J. & De Haan, E. H. F. The Corsi Block-Tapping Task: Standardization and normative data. Appl. Neuropsychol. 7, 252–258 (2000).

40. Rimland, B. & Edelson, S. Autism Treatment Evaluation Checklist. Autism Res. Inst.99 (1999).

41. Quezada, A. et al.. Usability Operations on Touch Mobile Devices for Users with Autism. J. Med. Syst. 41, 184 (2017).

42. De Vries, M., Prins, P. J. M., Schmand, B. A. & Geurts, H. M. Working memory and cognitive flexibility-training for children with an autism spectrum disorder: A randomized controlled trial. J. Child Psychol. Psychiatry Allied Discip. 56, 566–576 (2015).

43. Zinke, K. et al.. Visuospatial short-term memory explains deficits in tower task planning in high-functioning children with autism spectrum disorder. Child Neuropsychol. 16, 229–241 (2010).

